# Atrial Arrhythmia in Patients with Tetralogy of Fallot and Pulmonary Atresia with Intact Ventricular Septum: Risk of Thromboembolism and Bleeding

**DOI:** 10.1101/2025.05.21.25328122

**Authors:** Michael P O’Shea, Suganya Arunachalam Karikalan, Srekar Ravi, Matthew Van Ligten, Adam Bacon, Eiad Habib, Omar Baqal, Philip J Smyth, Winston Wang, Dani Green, Nneoma Alozie, Chelsea Marshal, Abhishek Deshmukh, Alexander Egbe, Heidi Connolly, Marlene Girardo, Hicham El Masry, David Majdalany

## Abstract

**Introduction:** Adults with congenital heart disease (CHD), particularly tetralogy of Fallot (ToF) and pulmonary atresia with intact ventricular septum (PA-IVS), are at increased risk for atrial fibrillation (AF) and its complications. We evaluated thromboembolic and bleeding risks in this population and assessed the predictive value of CHA_2_DS_2_-VASc and HAS-BLED scores.

**Methods:** In this retrospective cohort study, patients with ToF-PS, ToF-PA, or PA-IVS and AF were identified across Mayo Clinic sites (1999–2023) were analyzed. Primary outcomes were thromboembolism (stroke, TIA, systemic embolism) and major bleeding. Cox regression controlled for age; anticoagulation exposure was treated as time-varying. Risk score performance was evaluated using ROC curves.

**Results:** 300 patients with ToF-PS, ToF-PA, or PA-IVS were identified. Over 2,296 person-years, 12 thromboembolic events occurred (5.23/1000 person-years). Prior stroke, TIA, peripheral embolism, and Potts shunt were significant predictors. CHA_2_DS_2_-VASc had good discrimination (c-statistic 0.78); no events occurred in patients with a score of 0. Major bleeding occurred 69 times over 1,845 person-years (3.74/100 person-years), more frequently in ToF-PA (HR 2.05, p = 0.005), chronic AF, and those with Waterson shunts (HR 4.64, p = 0.013). DOAC use was associated with significantly higher bleeding risk than warfarin (HR 5.89, p < 0.001). The HAS-BLED score had limited predictive value (c-statistic 0.59) (figure 1).

**Discussion:** In CHD patients with AF, thromboembolic risk was low with anticoagulation, and CHA_2_DS_2_-VASc performed well. However, bleeding risk, particularly with DOACs and in patients with specific anatomical features, was high. Warfarin may be the safer anticoagulant in this population.

Figure 1:
Graphical Abstract

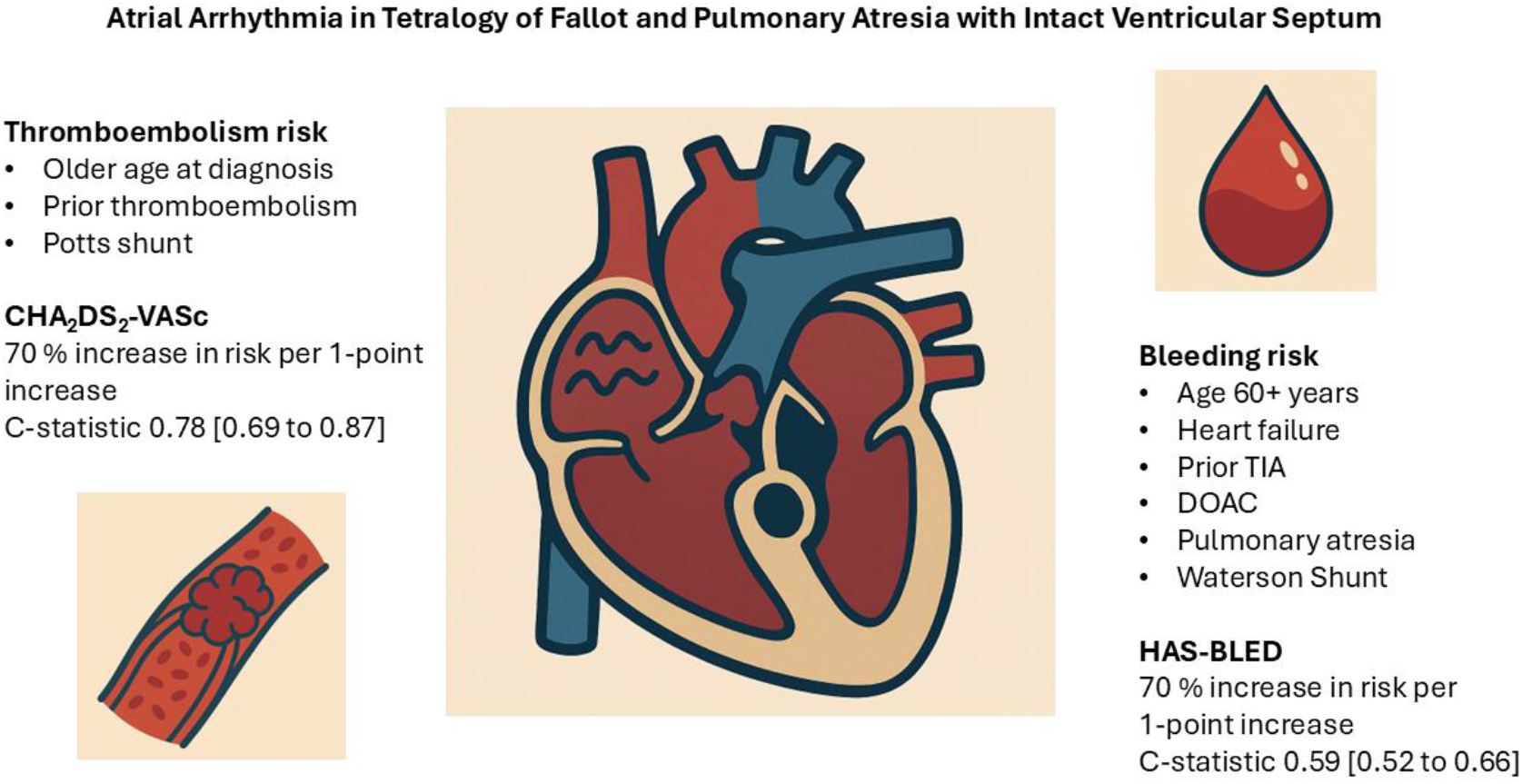

## Introduction

Advances in the care of complex congenital heart disease (CHD) have extended life expectancy into late adulthood(1-4). This has led to a growing need for clinical information on late complications of diseases such as tetralogy of Fallot (ToF) and pulmonary atresia with intact ventricular septum (PA-IVS)(5). Atrial fibrillation and flutter (AF) are common late complications of CHD repair, typically arising from surgical site substrate(6). AF in people with CHD is associated with increased rates of stroke, ventricular tachycardia and all-cause mortality(4, 7).

Tetralogy of Fallot (ToF) is the most common form of complex cyanotic CHD(8). It results from malalignment of the developing conal septum, causing an overriding aorta with right ventricular outflow tract obstruction (subpulmonic, pulmonic or suprapulmonic) with downstream sequelae including right ventricular hypertrophy and the development of ventricular septal defects(9).

There are two major sub-groups, ToF with Pulmonary Stenosis (ToF-PS) and ToF with Pulmonary Atresia (ToF-PA). Pulmonary Atresia with Intact Ventricular Septum (PA-IVS) is a form of complex CHD like ToF but without an overriding aorta or septal defect. These conditions are complicated by sequelae of right ventricular outflow tract obstruction with associated right ventricular hypertension, and are associated with a high prevalence of early onset atrial fibrillation(10-13). This study aims to explore the association between congenital disease-specific variables and major bleeding and stroke risk in people with ToF and PA-IVS who have atrial arrhythmias.

## Methods

Patients attending any Mayo Clinic site between January 1^st^ 1999 and October 1^st^ 2023 were included in this retrospective cohort study. Participants were included if they had a diagnosis of tetralogy of Fallot with pulmonic stenosis or pulmonic atresia, or pulmonic atresia with intact ventricular septum, and if they received treatment at the Mayo Clinic for atrial fibrillation or flutter.

Participants entered the study from onset of AF and exited the study after date of first heart transplant, time of death, time of last follow-up. When assessing anticoagulant effect, study entry time was adjusted to date of first anticoagulant initiation, and discontinuation was defined as cessation without re-initiation of the same anticoagulant in the subsequent 3 months. Participants were censored from event-specific analysis if they experienced the outcome being analyzed.

The primary outcomes were thromboembolic incidence and major bleeding incidence rates. Thromboembolic events were defined as a stroke, transient ischemic attack (TIA) or peripheral systemic arterial embolism. The International Society of Hemostasis and Thrombosis definition was used to determine major bleeding events(14).

Clinical variables were selected based on available information at the time of onset of AF. Where a variable was not available, a participant was excluded from analysis involving that specific variable. Missing data on baseline clinical variables was uncommon owing to comprehensive documentation, and did not exceed 5% for any individual variable.

Analysis was conducted using cox univariable regression, controlling for current age. For age category, lexis expansion was used to generate a time-varying variable. Receiver operator curve analysis was used to assess the performance of the CHA_2_DS_2_-VASc and HAS-BLED scores. Analysis was conducted using Stata IC 16. Graphical abstract was generated using ChatGPT (see supplemental appendix 1 for prompts used). Approval was obtained from the Mayo Clinic Institutional Review Board (ID 23-008909).

## Results

Three hundred participants were included in the study, of whom 161 (56.1%) had TOF-PS, 126 (46.9%) had TOF-PA, and 13 (4.3%) had PA-IVS. Two hundred and eighty-nine were followed longitudinally and were included in rate analysis, while all were retained for descriptive statistics.

### Thromboembolic events

There were 12 thromboembolic (TE) events observed over 2,296 person-years (mean follow-up 7.94 years), of which 10 were ischemic strokes, one was a transient ischemic attack, and one was a peripheral systemic arterial embolism. The rate of TE events was 5.23 [2.97 to 9.20] per 1000 person years. Comorbidities associated with TE events included prior stroke (HR 3.76 [95% CI 1.12 to 12.58], p=0.031) and TIA (HR 8.06 [95% CI 1.48 to 43.77], p=0.016), prior peripheral embolism (HR15.21 [95% CI 2.70 to 85.64, p=0.002), older age at diagnosis (p=0.045), and prior Potts (left pulmonary artery to descending aorta) shunt (HR 5.97 [95% CI 1.20 to 29.83], p=0.029) (Tables 1 & 2).

**Table 1:**
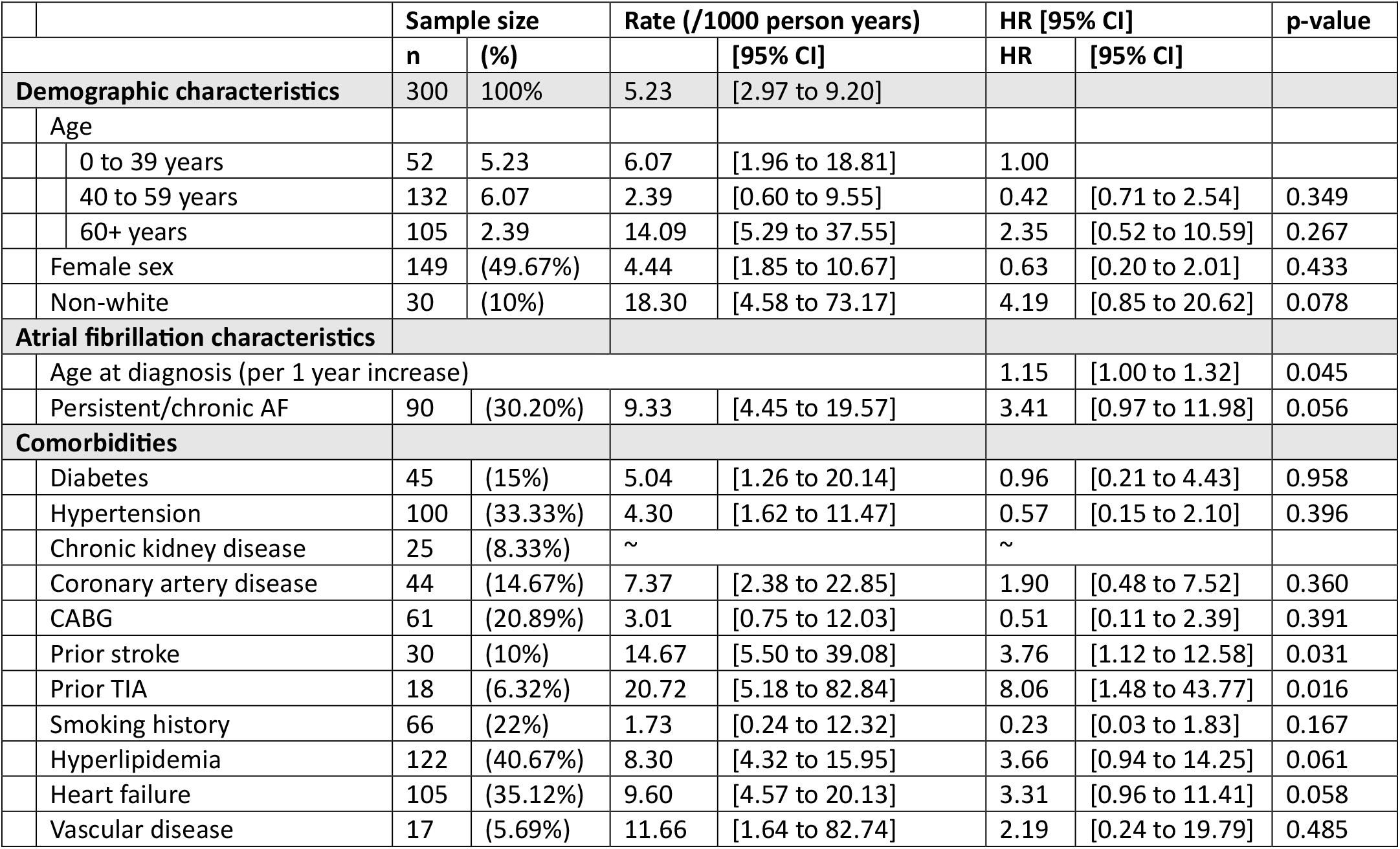
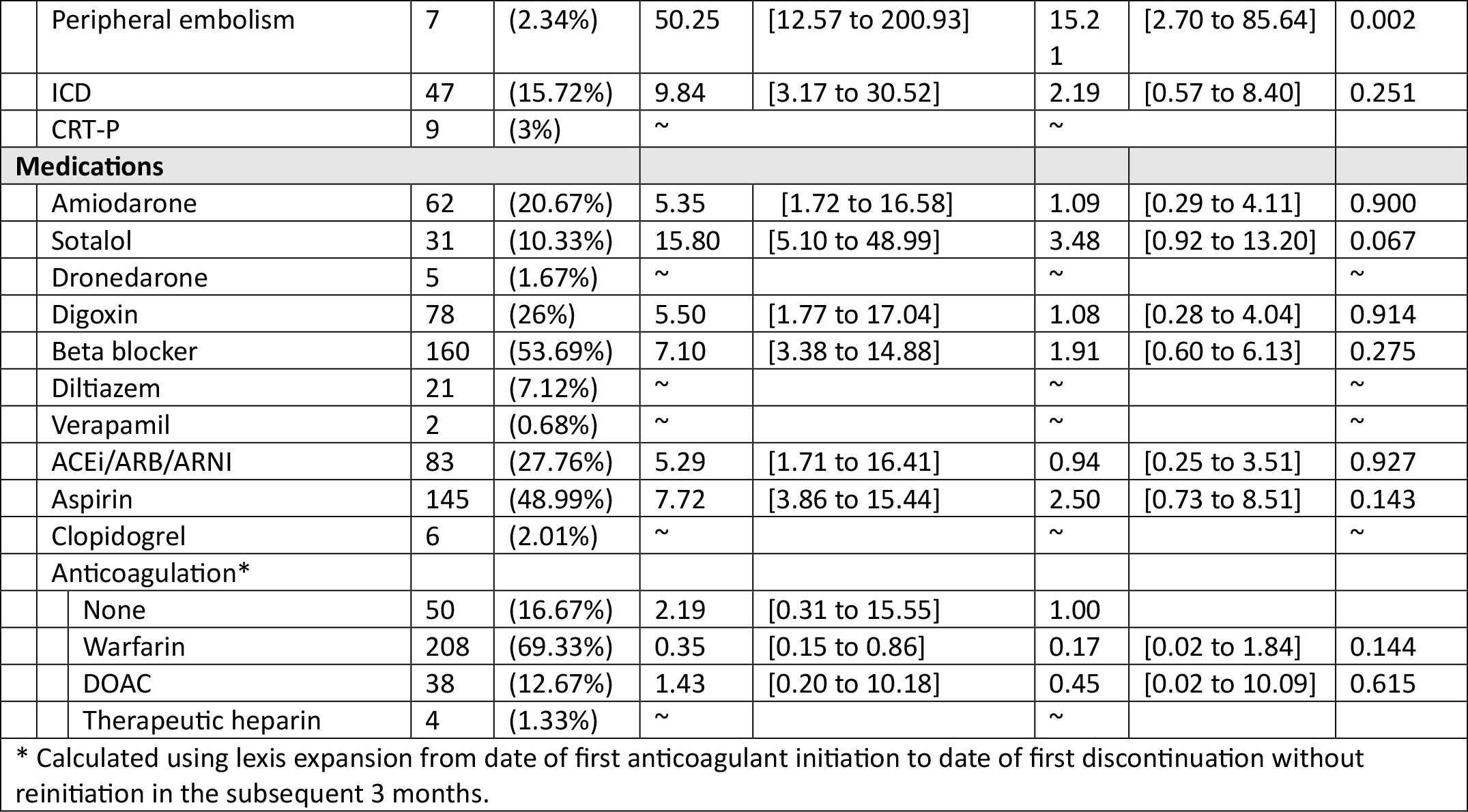
Characteristics and rate of thromboembolic events (stroke, TIA and peripheral embolism) among people with ToF-PS, ToF-PA and PA-IVS with concurrent atrial fibrillation, calculated using cox univariable regression controlling for current age.

|  | Sample size |  | Rate (/1000 person years) |  | HR [95% CI] |  | p-value |
| --- | --- | --- | --- | --- | --- | --- | --- |
|  | n | (%) |  | [95% CI] | HR | [95% CI] |  |
| **Demographic characteristics** | 300 | 100% | 5.23 | [2.97 to 9.20] |  |  |  |
| Age |  |  |  |  |  |  |  |
| 0 to 39 years | 52 | 5.23 | 6.07 | [1.96 to 18.81] | 1.00 |  |  |
| 40 to 59 years | 132 | 6.07 | 2.39 | [0.60 to 9.55] | 0.42 | [0.71 to 2.54] | 0.349 |
| 60+ years | 105 | 2.39 | 14.09 | [5.29 to 37.55] | 2.35 | [0.52 to 10.59] | 0.267 |
| Female sex | 149 | (49.67%) | 4.44 | [1.85 to 10.67] | 0.63 | [0.20 to 2.01] | 0.433 |
| Non-white | 30 | (10%) | 18.30 | [4.58 to 73.17] | 4.19 | [0.85 to 20.62] | 0.078 |
| **Atrial fibrillation characteristics** |  |  |  |  |  |  |  |
| Age at diagnosis (per 1 year increase) |  |  |  |  | 1.15 | [1.00 to 1.32] | 0.045 |
| Persistent/chronic AF | 90 | (30.20%) | 9.33 | [4.45 to 19.57] | 3.41 | [0.97 to 11.98] | 0.056 |
| **Comorbidities** |  |  |  |  |  |  |  |
| Diabetes | 45 | (15%) | 5.04 | [1.26 to 20.14] | 0.96 | [0.21 to 4.43] | 0.958 |
| Hypertension | 100 | (33.33%) | 4.30 | [1.62 to 11.47] | 0.57 | [0.15 to 2.10] | 0.396 |
| Chronic kidney disease | 25 | (8.33%) | ~ |  | ~ |  |  |
| Coronary artery disease | 44 | (14.67%) | 7.37 | [2.38 to 22.85] | 1.90 | [0.48 to 7.52] | 0.360 |
| CABG | 61 | (20.89%) | 3.01 | [0.75 to 12.03] | 0.51 | [0.11 to 2.39] | 0.391 |
| Prior stroke | 30 | (10%) | 14.67 | [5.50 to 39.08] | 3.76 | [1.12 to 12.58] | 0.031 |
| Prior TIA | 18 | (6.32%) | 20.72 | [5.18 to 82.84] | 8.06 | [1.48 to 43.77] | 0.016 |
| Smoking history | 66 | (22%) | 1.73 | [0.24 to 12.32] | 0.23 | [0.03 to 1.83] | 0.167 |
| Hyperlipidemia | 122 | (40.67%) | 8.30 | [4.32 to 15.95] | 3.66 | [0.94 to 14.25] | 0.061 |
| Heart failure | 105 | (35.12%) | 9.60 | [4.57 to 20.13] | 3.31 | [0.96 to 11.41] | 0.058 |
| Vascular disease | 17 | (5.69%) | 11.66 | [1.64 to 82.74] | 2.19 | [0.24 to 19.79] | 0.485 |

|  |  |  |  |  |  |  |  |  |
| --- | --- | --- | --- | --- | --- | --- | --- | --- |
|  | Peripheral embolism | 7 | (2.34%) | 50.25 | [12.57 to 200.93] | 15.2<br>1 | [2.70 to 85.64] | 0.002 |
|  | ICD | 47 | (15.72%) | 9.84 | [3.17 to 30.52] | 2.19 | [0.57 to 8.40] | 0.251 |
|  | CRT-P | 9 | (3%) | ~ |  | ~ |  |  |
| **Medications** |  |  |  |  |  |  |  |  |
|  | Amiodarone | 62 | (20.67%) | 5.35 | [1.72 to 16.58] | 1.09 | [0.29 to 4.11] | 0.900 |
|  | Sotalol | 31 | (10.33%) | 15.80 | [5.10 to 48.99] | 3.48 | [0.92 to 13.20] | 0.067 |
|  | Dronedarone | 5 | (1.67%) | ~ |  | ~ |  | ~ |
|  | Digoxin | 78 | (26%) | 5.50 | [1.77 to 17.04] | 1.08 | [0.28 to 4.04] | 0.914 |
|  | Beta blocker | 160 | (53.69%) | 7.10 | [3.38 to 14.88] | 1.91 | [0.60 to 6.13] | 0.275 |
|  | Diltiazem | 21 | (7.12%) | ~ |  | ~ |  | ~ |
|  | Verapamil | 2 | (0.68%) | ~ |  | ~ |  | ~ |
|  | ACEi/ARB/ARNI | 83 | (27.76%) | 5.29 | [1.71 to 16.41] | 0.94 | [0.25 to 3.51] | 0.927 |
|  | Aspirin | 145 | (48.99%) | 7.72 | [3.86 to 15.44] | 2.50 | [0.73 to 8.51] | 0.143 |
|  | Clopidogrel | 6 | (2.01%) | ~ |  | ~ |  | ~ |
|  | Anticoagulation* |  |  |  |  |  |  |  |
|  | None | 50 | (16.67%) | 2.19 | [0.31 to 15.55] | 1.00 |  |  |
|  | Warfarin | 208 | (69.33%) | 0.35 | [0.15 to 0.86] | 0.17 | [0.02 to 1.84] | 0.144 |
|  | DOAC | 38 | (12.67%) | 1.43 | [0.20 to 10.18] | 0.45 | [0.02 to 10.09] | 0.615 |
|  | Therapeutic heparin | 4 | (1.33%) | ~ |  | ~ |  |  |
| * Calculated using lexis expansion from date of first anticoagulant initiation to date of first discontinuation without reinitiation in the subsequent 3 months. |  |  |  |  |  |  |  |  |

**Table 2:** Congenital heart disease-specific variables and rate of thromboembolic events (stroke, TIA and peripheral embolism) among people with ToF-PS, ToF-PA and PA-IVS with concurrent atrial fibrillation, controlling for current age.

|  |  |  |  |  |  |  |  |  |
| --- | --- | --- | --- | --- | --- | --- | --- | --- |
|  |  | **Sample size** |  | **Rate (/1000 person years)** |  | **Hazard ratio** |  | **p-value** |
|  |  | **n** | **(%)** |  | **[95% CI]** | **HR** | **[95% CI]** |  |
| **Subtype** |  |  |  |  |  |  |  |  |
|  | TOF-PS | 161 | (53.67%) | 4.13 | [1.86 to 9.20] | 1.00 |  |  |
|  | TOF-PA | 126 | (95.67%) | 8.73 | [3.92 to 19.44] | 2.30 | [0.71 to 7.46] | 0.164 |
|  | PA-IVS | 13 | (4.33%) | ~ |  | ~ |  |  |
| **Shunt types** |  |  |  |  |  |  |  |  |
|  | Any shunt | 154 | (51.33%) | 5.49 | [2.62 to 11.52] | 1.08 | [0.34 to 3.47] | 0.893 |
|  | Atrial shunt | 23 | (8.98%) | 16.91 | [4.23 to 67.61] | 3.32 | [0.69 to 16.00] | 0.135 |
|  | BT shunt | 92 | (30.67%) | 4.27 | [1.38 to 13.25] | 0.82 | [0.22 to 3.06] | 0.768 |
|  | Glenn shunt | 2 | (0.67%) | ~ |  | ~ |  |  |
|  | Central shunt | 51 | (17%) | 4.56 | [1.14 to 18.25] | 0.84 | [0.17 to 4.01] | 0.823 |
|  | Balloon valvotomy | 34 | (11.33%) | ~ |  | ~ |  |  |
|  | Potts shunt | 8 | (2.67%) | 33.78 | [8.45 to 135.06] | 5.97 | [1.20 to 29.83] | 0.029 |
|  | Waterson shunt | 9 | (3%) | ~ |  | ~ |  |  |
|  | Unifocalization | 13 | (4.33%) | 10.43 | [1.47 to 74.07] | 2.97 | [0.35 to 25.19] | 0.318 |
| **Definitive repair** |  |  |  |  |  |  |  |  |
|  | TAP repair | 145 | (48.33%) | 5.60 | [2.52 to 12.47] | 1.06 | [0.34 to 3.34] | 0.924 |
|  | Non-TAP repair | 148 | (49.33%) | 4.96 | [2.23 to 11.03] | 0.92 | [0.29 to 2.87] | 0.882 |
|  | RV-PA conduit | 78 | (26%) | 7.33 | [2.75 to 19.53] | 1.72 | [0.51 to 5.86] | 0.385 |
|  | Repair with PVR | 174 | (58.19%) | 5.85 | [2.92 to 11.69] | 1.68 | [0.50 to 5.71] | 0.405 |
| **Valve replacement** |  |  |  |  |  |  |  |  |
|  | Bioprosthetic valve | 125 | (41.67%) | 7.12 | [3.20 to 15.85] | 1.72 | [0.54 to 5.95] | 0.342 |
|  | Metallic valve | 18 | (6%) | 7.77 | [1.09 to 55.17] | 2.51 | [0.28 to 22.48] | 0.411 |

The CHA_2_DS_2_-VASc score was associated with increased TE events (p=0.044) and had a strong predictive value across the whole cohort (c-statistic 0.78 [95% CI 0.69 to 0.87]) (figure 2). There were no major thromboembolic events among people with a CHA_2_DS_2_-VASc score of 0 (over 319.7 person years observed), and 2.70 [95% CI 0.68 to 10.81] events per 1000 person years for those with a CHA_2_DS_2_-VASc score of 1 (Table 3). There was weak evidence of departure from a linear trend (LR χ^2^ (3) = 7.39, p=0.060) for scores 1-5. For each 1-point increase in CHA_2_DS_2_-VASc score, there was an additional 1.73% [95% CI 1.14 to 2.26] increase in the rate of TE events (p=0.010) when linearity was assumed.

**Table 3:** Rate of thromboembolic events among people with congenital heart disease and atrial fibrillation, stratified by CHA_2_DS_2_-VASc score.

| CHA<sub>2</sub>DS<sub>2</sub>-VASc score | n | Events | Person time | Stroke Rate | HR [95% CI] | p-value |
| --- | --- | --- | --- | --- | --- | --- |
|  |  |  | (years) | (per 1000 person years) |  |  |
| 0 | 54 | 0 | 319.7 | 0 |  |  |
| 1 | 98 | 2 | 740 | 2.70 [0.68 to 10.81] | 1.00 |  |
| 2 | 84 | 2 | 704 | 2.84 [0.71 to 11.36] | 1.05 [0.148 to 7.46] | 0.960 |
| 3 | 42 | 6 | 287 | 20.91 [9.39 to 46.54] | 7.74 [1.56 to 38.33] | 0.003 |
| 4 | 21 | 1 | 181.4 | 5.51 [0.78 to 39.12] | 2.04 [0.19 to 22.49] | 0.552 |
| 5 | 11 | 1 | 63.9 | 15.65 [2.21 to ~] | 5.79 [0.53 to 63.88] | 0.104 |
| C-statistic |  |  |  | 0.78 [0.69 to 0.87] |  |  |
| Test for departure from linearity |  |  |  | LR Chi (3) = 7.39, p=0.060 |  |  |
| Per 1 point increase assuming linear trend for scores 1-5 |  |  |  |  | 1.73 [1.14 to 2.62] | 0.010 |

**Figure 2:**
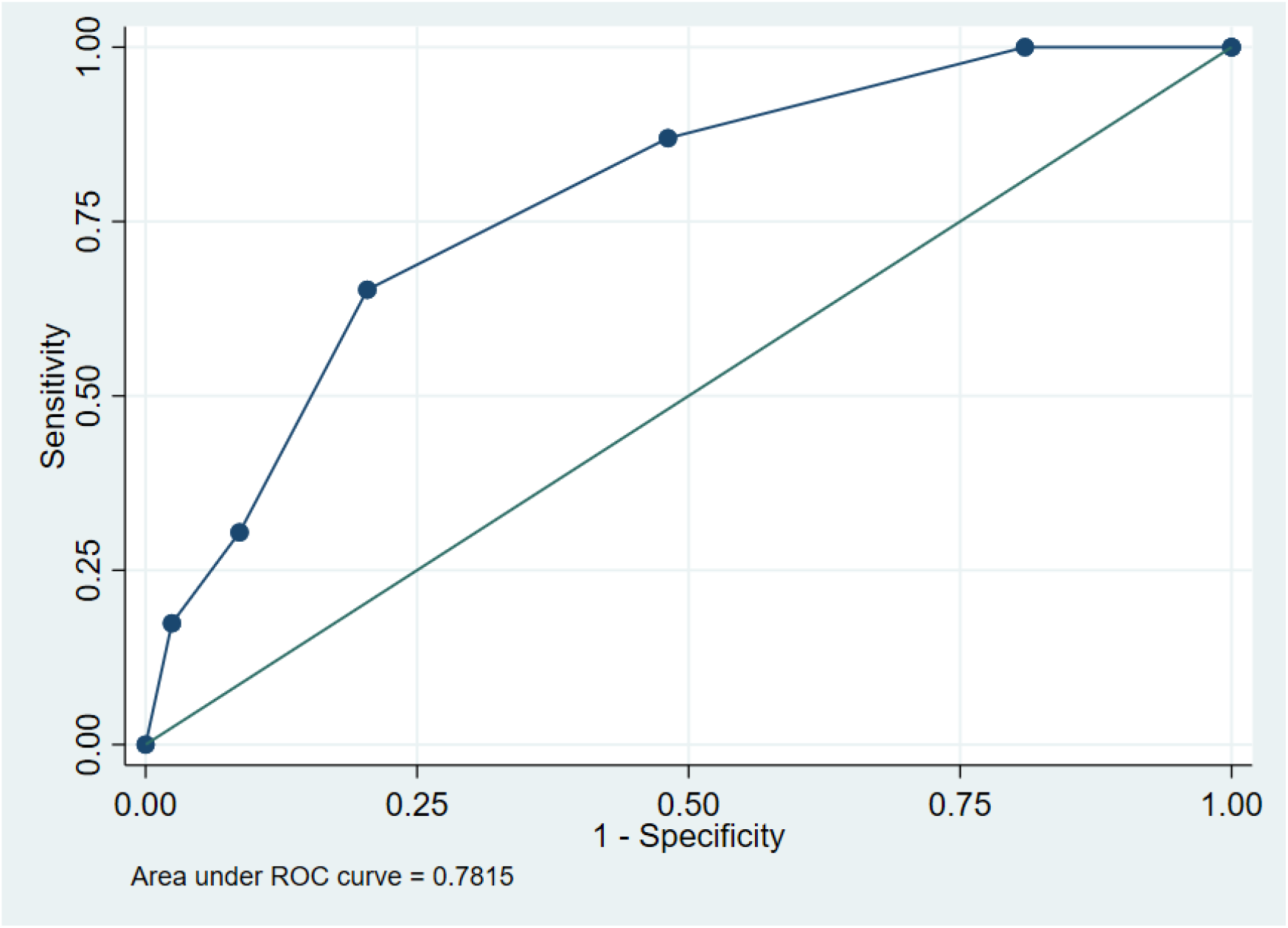
Receiver operator curve of the CHA_2_DS_2_-VASc score in predicting thromboembolic events in a cohort of patients with AF with ToF and PA-IVS.

### Major bleeding events

There were 69 major bleeding events over 1,844.98 person years (mean follow-up 6.38 years). The bleeding rate was 3.74 [2.95 to 4.74] events per 100 person years. Major bleeding was more than twice as common among non-white people (p=0.018) and among those age 60 and older (p=0.030) (Table 4). Chronic atrial fibrillation (p=0.02), older age at diagnosis (p=0.027), history of coronary artery bypass grafting (p=0.002), heart failure (p=0.005), and history of peripheral embolism (p=0.029) were all associated with higher bleeding rates. The most common anticoagulant used was warfarin (208). Among those using DOACs, 26 used apixaban, 9 used rivaroxaban and 3 used edoxaban. Major bleeding events were 5 times more common among those taking direct oral anticoagulants compared to those using warfarin (HR 5.89 [2.81 to 12.35], p<0.001).

**Table 4:**
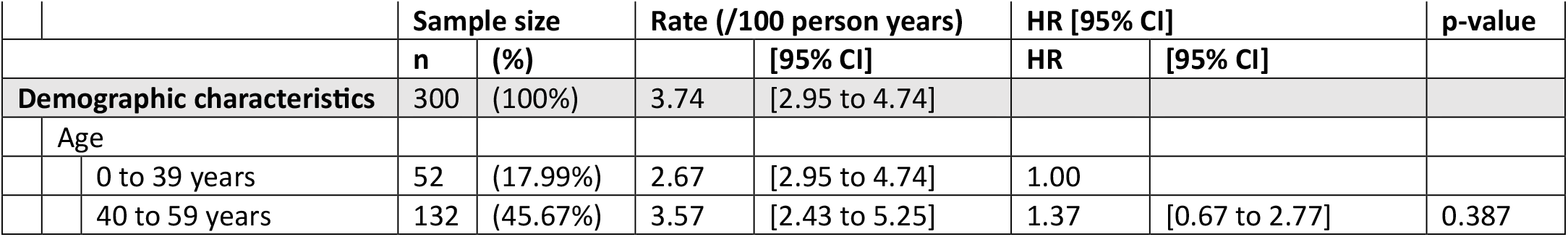
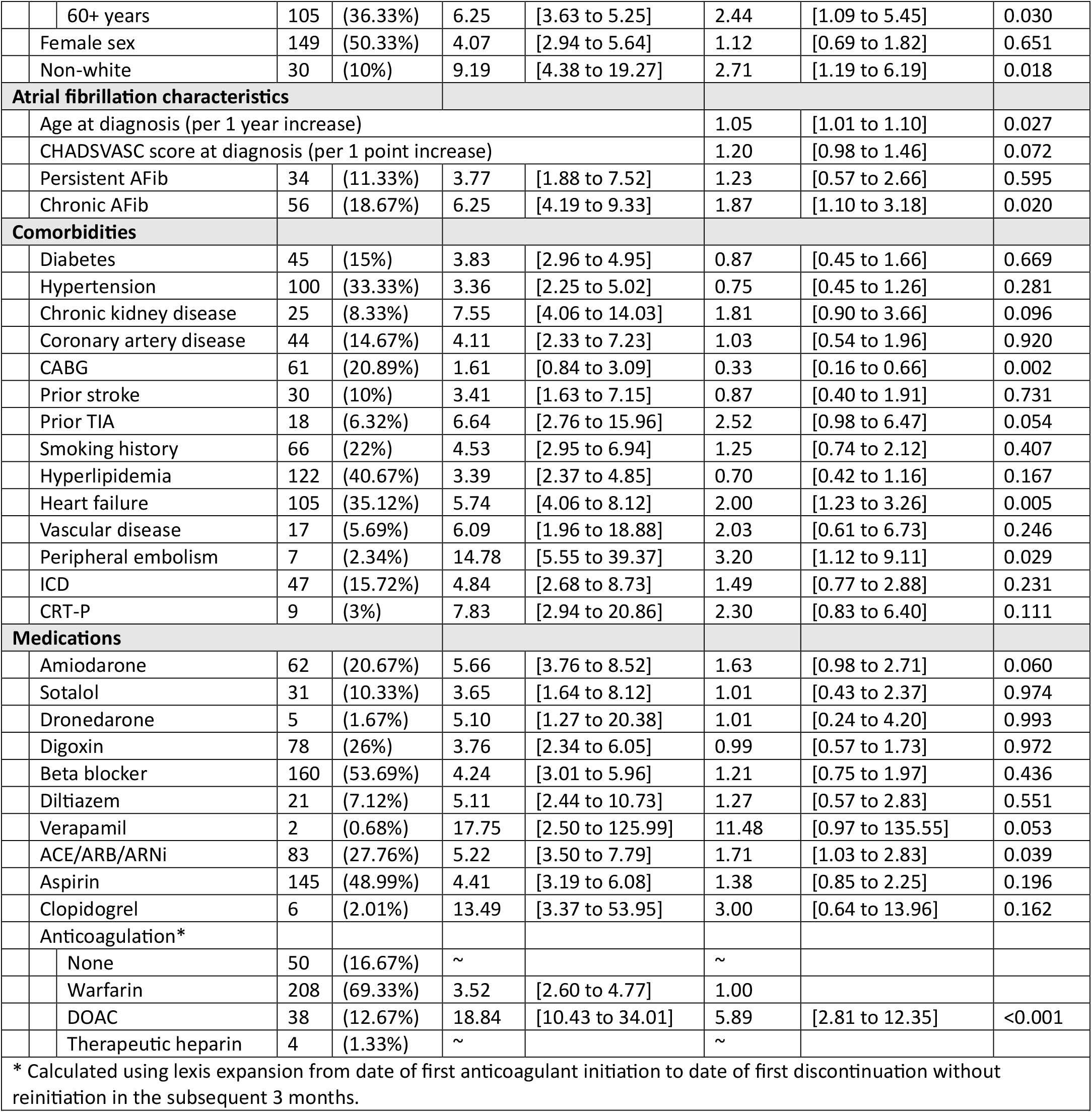
Characteristics and rate of major bleeding events among people with ToF-PS, ToF-PA and PA-IVS with concurrent atrial fibrillation, calculated using univariable cox regression controlling for current age.

There were twice as many major bleeding events associated with TOF-PA compared to TOF-PS (HR =2.05 [1.24 to 3.38], p=0.005). There was a four-fold increase in major bleeding events among people who previously underwent a Waterson shunt (right pulmonary artery to ascending aorta) (HR = 4.64, p=0.013) (Table 5).

**Table 5:** Congenital heart disease-specific variables and rate of major bleeding among people with ToF-PS, ToF-PA and PA-IVS with concurrent atrial fibrillation, controlling for current age.

|  |  | Sample size |  | Rate (/1000 person years) |  | Hazard ratio |  | p-value |
| --- | --- | --- | --- | --- | --- | --- | --- | --- |
|  |  | n | (%) |  | [95% CI] | HR | [95% CI] |  |
| **Subtype** |  |  |  |  |  |  |  |  |
|  | TOF-PS | 161 | (53.67%) | 2.92 | [2.09 to 4.09] | 1.00 |  |  |
|  | TOF-PA | 126 | (95.67%) | 5.40 | [3.79 to 7.67] | 2.05 | [1.24 to 3.38] | 0.005 |
|  | PA-IVS | 13 | (4.33%) | 3.73 | [1.40 to 9.95] | 1.77 | [0.50 to 6.22] | 0.375 |
| **Shunt types** |  |  |  |  |  |  |  |  |
|  | Any shunt | 154 | (51.33%) | 3.83 | [2.80 to 5.25] | 1.15 | [0.70 to 1.87] | 0.583 |
|  | Atrial shunt | 23 | (8.98%) | 3.22 | [1.04 to 9.98] | 0.73 | [0.22 to 2.42] | 0.604 |
|  | BT shunt | 92 | (30.67%) | 4.55 | [3.05 to 6.80] | 1.49 | [0.89 to 2.48] | 0.130 |
|  | Glenn shunt | 2 | (0.67%) | 5.01 | [0.71 to 35.58] | 1.14 | [0.14 to 9.57] | 0.901 |
|  | Central shunt | 51 | (17%) | 4.95 | [3.08 to 7.96] | 1.56 | [0.88 to 2.74] | 0.126 |
|  | Balloon valvotomy | 34 | (11.33%) | 2.60 | [1.17 to 5.80] | 0.68 | [0.29 to 1.59] | 0.374 |
|  | Potts shunt | 8 | (2.67%) | 3.75 | [0.94 to 14.98] | 0.90 | [0.21 to 3.78] | 0.881 |
|  | Waterson shunt | 9 | (3%) | 14.13 | [4.56 to 43.80] | 4.64 | [1.38 to 15.63] | 0.013 |
|  | Unifocalization | 13 | (4.33%) | 4.55 | [1.71 to 12.13] | 1.28 | [0.45 to 3.58] | 0.644 |
| **Definitive repair** |  |  |  |  |  |  |  |  |
|  | TAP repair | 145 | (48.33%) | 4.36 | [3.19 to 5.97] | 1.45 | [0.89 to 2.35] | 0.137 |
|  | Non-TAP repair | 148 | (49.33%) | 3.71 | [2.67 to 5.17] | 0.94 | [0.58 to 1.53] | 0.815 |
|  | RV-PA conduit | 78 | (26%) | 3.65 | [2.30 to 5.79] | 1.02 | [0.59 to 1.77] | 0.938 |
|  | Repair with PVR | 174 | (58.19%) | 4.08 | [3.05 to 5.44] | 1.24 | [0.75 to 2.06] | 0.404 |
| **Valve replacement** |  |  |  |  |  |  |  |  |
|  | Bioprosthetic valve | 125 | (41.67%) | 4.48 | [3.13 to 6.41] | 1.55 | [0.94 to 2.57] | 0.085 |
|  | Metallic valve | 18 | (6%) | 5.75 | [3.13 to 6.41] | 1.82 | [0.75 to 4.42] | 0.183 |

The HAS-BLED score performed only slightly better than chance in this cohort, with a score range of 0 to 3 (receiver operator curve (ROC) area = 0.59 [0.52 to 0.66]) (figure 3). The rate of major bleeding events among people with a HAS-BLED score of 0 was 2.61 [95% CI 1.54 to 4.40] per 100 person years.

**Table 6:**
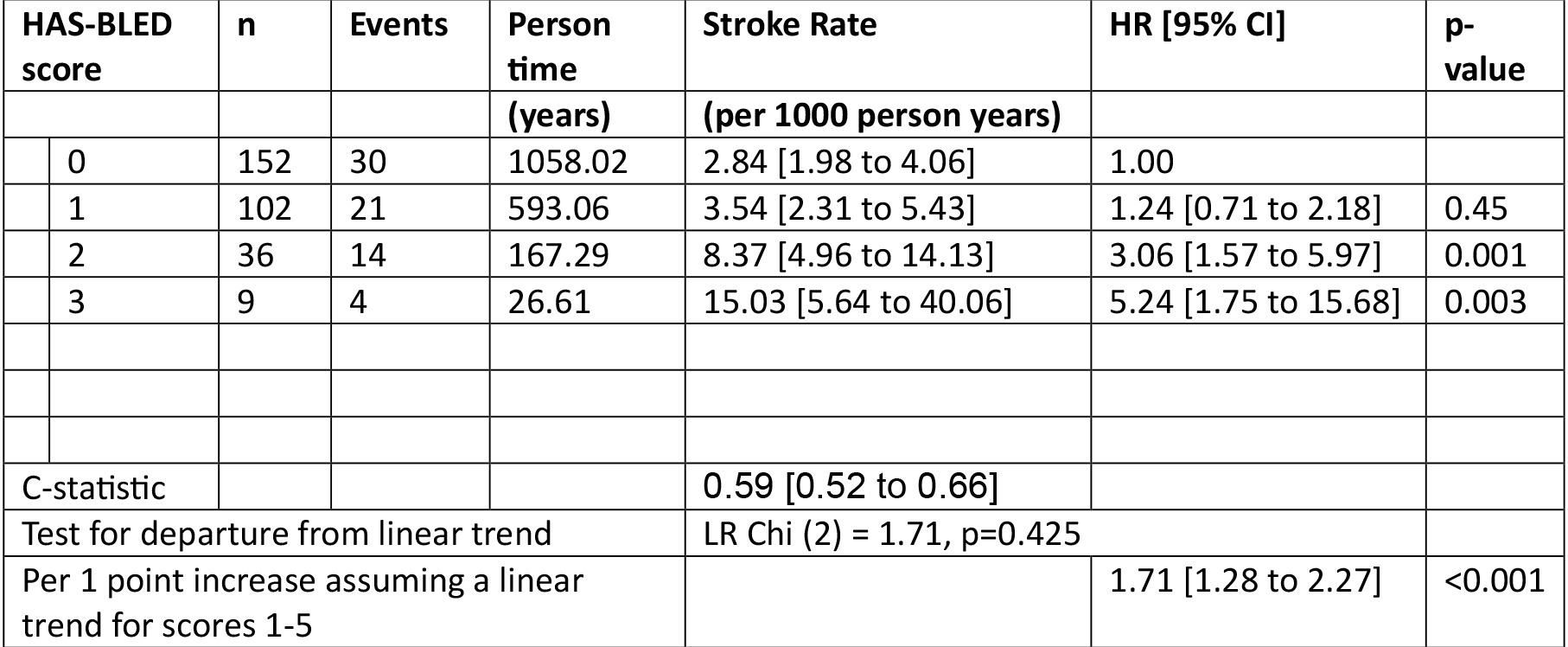
Rate of major bleeding events among people with congenital heart disease and atrial fibrillation, stratified by HAS-BLED score.

|  |  |  |  |  |  |  |
| --- | --- | --- | --- | --- | --- | --- |
| HAS-BLED score | n | Events | Person time | Stroke Rate | HR [95% CI] | p-value |
|  |  |  | (years) | (per 1000 person years) |  |  |
| 0 | 152 | 30 | 1058.02 | 2.84 [1.98 to 4.06] | 1.00 |  |
| 1 | 102 | 21 | 593.06 | 3.54 [2.31 to 5.43] | 1.24 [0.71 to 2.18] | 0.45 |
| 2 | 36 | 14 | 167.29 | 8.37 [4.96 to 14.13] | 3.06 [1.57 to 5.97] | 0.001 |
| 3 | 9 | 4 | 26.61 | 15.03 [5.64 to 40.06] | 5.24 [1.75 to 15.68] | 0.003 |
| C-statistic |  |  |  | 0.59 [0.52 to 0.66] |  |  |
| Test for departure from linear trend |  |  |  | LR Chi (2) = 1.71, p=0.425 |  |  |
| Per 1 point increase assuming a linear trend for scores 1-5 |  |  |  |  | 1.71 [1.28 to 2.27] | <0.001 |

**Figure 3:**
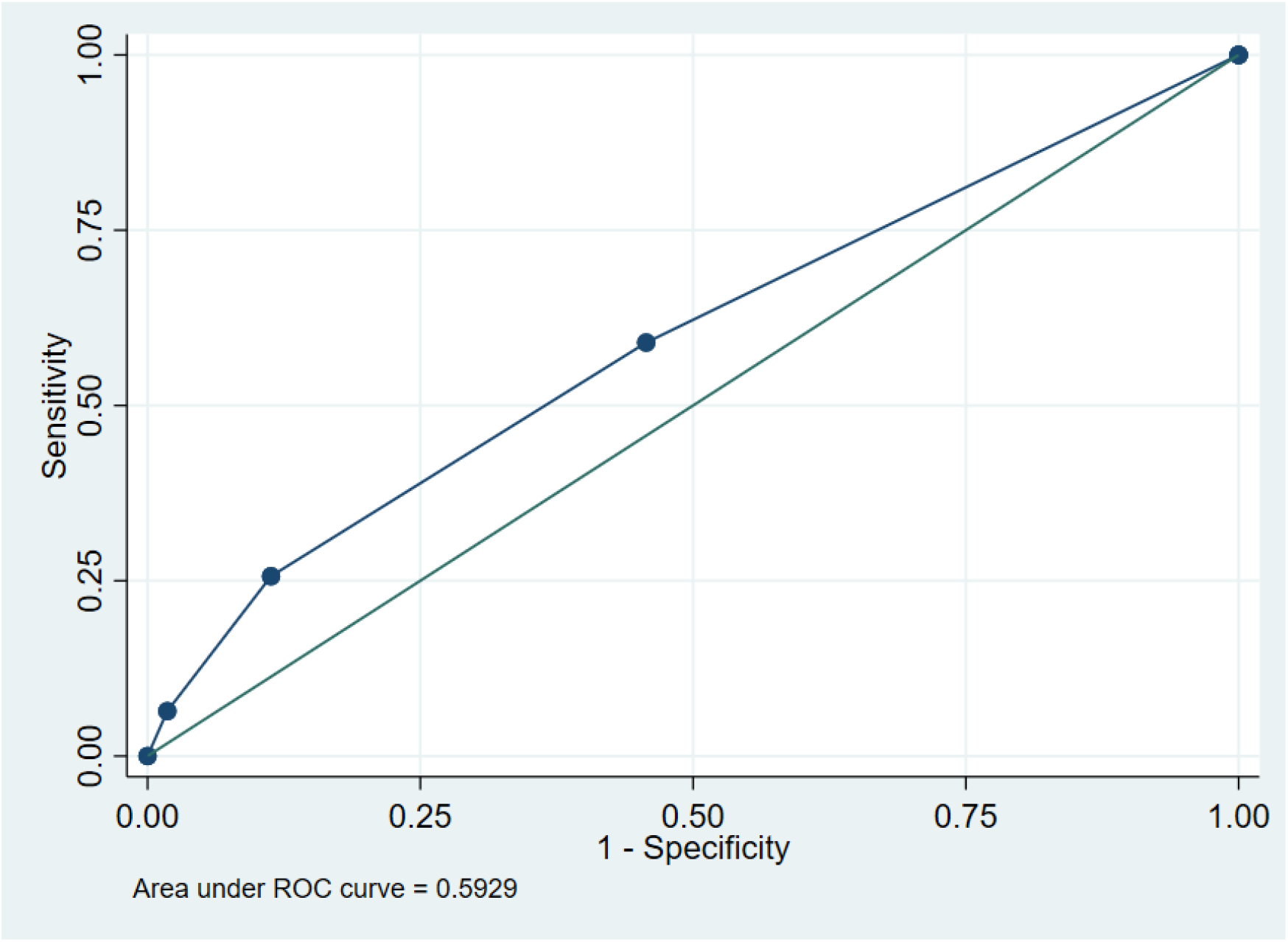
Receiver operator curve of the HAS-BLED score in predicting major bleeding events in a cohort of patients with AF with ToF and PA-IVS.

## Discussion

This study identified a 5-fold increased risk of bleeding among people with ToF and PA-IVS who developed AF who received a DOAC compared to those who received warfarin. This has not been previously reported, likely due to the complexity-based stratification of CHD in other studies. The TACTIC study demonstrated that high complexity CHD was associated with increased rates of major bleeding in people with AF, however, it did not differentiate between CHD subgroups, nor did it present results exploring differences in bleeding outcomes in warfarin compared to DOACs(7). Subsequent studies exploring DOAC safety in CHD patients did not examine differences in major bleeding event rates by CHD subtypes beyond complexity level(15-20) or unifocalization(21).

It is possible that subtypes of CHD associated with right ventricular hypertension have a higher bleeding risk compared to subtypes that do not. In this study, people who had anatomical features associated with higher right ventricular afterload states, in particular Waterston Shunts and pulmonary atresia rather than pulmonary stenosis, had higher rates of major bleeding. We previously demonstrated that elevated right ventricular systolic pressure (RVSP) was associated with excess bleeding with certain DOACs in non-CHD adults with atrial fibrillation(22, 23).

Mechanisms by which this could cause excess bleeding include the synergistic effect of elevated central venous pressure and site-specific gastrointestinal bleeding risk of DOACs and altered drug-protein binding with congestive hepatopathy.

The CHA_2_DS_2_-VASc score was an effective tool in predicting thromboembolic events. The rate of TE was low overall, likely due to the high use of anticoagulation in this cohort. Prior stroke, prior TIA and CABG, may be associated with TE through the association with atheromatous disease with atheromatous ischemic infarction rather than embolic stroke(24, 25). The HAS-BLED score had similar predictive value to the general population(26, 27).

The bleeding and stroke rates observed in this study are similar to those seen in other studies of complex CHD patients(7, 16, 19). Several other associations were seen, including increasing thromboembolic events with age in this relatively young cohort and (1).

Limitations of this study include the relatively small number of people taking specific DOACs, which prevented DOAC-specific sub-group analysis. This study involved an extensive exploration of disease-specific predictors of two AF outcomes. As such the risk of inadvertently assuming an association is present when it is not must be weighed against the biologic plausibility of the explanation until external validation or prospective. Multivariable regression was not performed due to data sparsity, with low prevalence of many binary variables as well as low overall event rates.

This study has several clinical implications. The identification of higher bleeding rates in people with ToF and PA-IVS using DOACs suggests warfarin may be a safer anticoagulant choice for these patients. The CHA_2_DS_2_-VASc score provided a robust estimate of thromboembolic events in this predominantly anticoagulated cohort, indicating its clinical utility in ToF and PA-IVS. These results reaffirm the limited value of the HAS-BLED score in predicting major bleeding events (28, 29). Several disease-specific risk factors for major bleeding were identified (including pulmonary atresia and prior Waterson shunt). While additional research is needed to determine the optimal thromboembolism risk-reduction strategy in this cohort, the results presented can help inform more accurate risk benefit discussions with these high bleeding-risk individuals.

Larger multicenter studies are needed to explore the difference in bleeding rates between different anticoagulants in CHD associated with right ventricular hypertension. Future research could explore different sub-groups of CHD who share similar physiology, including people with pulmonary valve stenosis without the tetrad of ToF and double chamber right ventricle.

Echocardiographic or hemodynamic evidence of a biologic gradient between right ventricle pressures or function and major bleeding events could better contextualize the associations observed in this study.

## Conclusion

In patients with PA-IVS and ToF with AF, DOAC use was associated with a five-fold increase in major bleeding events compared with warfarin. People with anatomic features associated with high right ventricular afterload, such as Waterson Shunts and Pulmonary Atresia, had higher bleeding rates. The CHA_2_DS_2_-VASc score was an effective tool in predicting thromboembolic events, while the HAS-BLED score performed no better than chance in predicting major bleeding events.

## Data Availability

The data used in this manuscript will be available in an irrevocably anonymized format on reasonable request submitted to the corresponding author within 12 months of publication.

## ABBREVIATIONS

CHD: Congenital Heart Disease
ToF: Tetralogy of Fallot
ToF-PA: Tetralogy of Fallot with Pulmonary Atresia
ToF-PS: Tetralogy of Fallot with Pulmonary Stenosis
PA-IVS: Pulmonary Atresia with Intact Ventricular Septum
AF: refers to both Atrial fibrillation and Atrial flutter (used interchangeably)
TIA: Transient Ischemic Attack
DOAC: Direct Oral Anticoagulant
TE: Thromboembolic Events
CABG: Coronary Artery Bypass Grafting
ICD: Implanted Cardiac Defibrillator
CRT-P: Cardiac Resynchronization Therapy Pacemaker
ACEi: Angiotensin Converting Enzyme Inhibitor
ARB: Angiotensin Receptor Blocker
ARNI: Angiotensin receptor neprilysin inhibitor
BT shunt: Blalock Taussig Shunt
TAP: Transannular patch
PVR: Pulmonary valve replacement
RVSP: Right ventricular systolic pressure
COPD: Chronic obstructive lung disease

## References

1. Tsui C, Wan D, Grewal J, Kiess M, Barlow A, Human D, et al. Increasing age and atrial arrhythmias are associated with increased thromboembolic events in a young cohort of adults with repaired tetralogy of Fallot. J Arrhythm. 2021;37(6):1546–54.

2. Lillehei CW, Cohen M, Warden HE, Read RC, Aust JB, Dewall RA, et al. Direct vision intracardiac surgical correction of the tetralogy of Fallot, pentalogy of Fallot, and pulmonary atresia defects; report of first ten cases. Ann Surg. 1955;142(3):418–42.

3. Marelli AJ, Ionescu-Ittu R, Mackie AS, Guo L, Dendukuri N, Kaouache M. Lifetime prevalence of congenital heart disease in the general population from 2000 to 2010. Circulation. 2014;130(9):749–56.

4. Possner M, Tseng SY, Alahdab F, Bokma JP, Lubert AM, Khairy P, et al. Risk Factors for Mortality and Ventricular Tachycardia in Patients With Repaired Tetralogy of Fallot: A Systematic Review and Meta-analysis. Canadian Journal of Cardiology. 2020;36(11):1815–25.

5. Baumgartner H, De Backer J, Babu-Narayan SV, Budts W, Chessa M, Diller G-P, et al. 2020 ESC Guidelines for the management of adult congenital heart disease: The Task Force for the management of adult congenital heart disease of the European Society of Cardiology (ESC). Endorsed by: Association for European Paediatric and Congenital Cardiology (AEPC), International Society for Adult Congenital Heart Disease (ISACHD). European Heart Journal. 2020;42(6):563–645.

6. de Groot NM, Lukac P, Schalij MJ, Makowski K, Szili-Torok T, Jordaens L, et al. Long-term outcome of ablative therapy of post-operative atrial tachyarrhythmias in patients with tetralogy of Fallot: a European multi-centre study. Europace. 2012;14(4):522–7.

7. Khairy P, Aboulhosn J, Broberg CS, Cohen S, Cook S, Dore A, et al. Thromboprophylaxis for atrial arrhythmias in congenital heart disease: A multicenter study. Int J Cardiol. 2016;223:729–35.

8. Apitz C, Webb GD, Redington AN. Tetralogy of Fallot. The Lancet. 2009;374(9699):1462–71.

9. Becker AE, Connor M, Anderson RH. Tetralogy of Fallot: a morphometric and geometric study. Am J Cardiol. 1975;35(3):402–12.

10. Reller MD, Strickland MJ, Riehle-Colarusso T, Mahle WT, Correa A. Prevalence of congenital heart defects in metropolitan Atlanta, 1998-2005. J Pediatr. 2008;153(6):807–13.

11. Khairy P, Aboulhosn J, Gurvitz MZ, Opotowsky AR, Mongeon FP, Kay J, et al. Arrhythmia burden in adults with surgically repaired tetralogy of Fallot: a multi-institutional study. Circulation. 2010;122(9):868–75.

12. Egbe AC, Vallabhajosyula S, Vojjini R, Banala K, Najam M, Faizee F, et al. Prevalence and in-hospital mortality during arrhythmia-related admissions in adults with tetralogy of Fallot. Int J Cardiol. 2019;297:49–54.

13. Wu MH, Chiu SN, Tseng WC, Lu CW, Kao FY, Huang SK. Atrial fibrillation in adult congenital heart disease and the general population. Heart Rhythm. 2023;20(9):1248–54.

14. Schulman S, Kearon C. Definition of major bleeding in clinical investigations of antihemostatic medicinal products in non-surgical patients. Journal of Thrombosis and Haemostasis. 2005;3(4):692–4.

15. Pujol C, Müssigmann M, Schiele S, Nagdyman N, Niesert A-C, Kaemmerer H, et al. Direct oral anticoagulants in adults with congenital heart disease - a single centre study. International Journal of Cardiology. 2020;300:127–31.

16. Yang H, Bouma BJ, Dimopoulos K, Khairy P, Ladouceur M, Niwa K, et al. Non-vitamin K antagonist oral anticoagulants (NOACs) for thromboembolic prevention, are they safe in congenital heart disease? Results of a worldwide study. Int J Cardiol. 2020;299:123–30.

17. Brás PG, Mano TB, Rito T, Castelo A, Ferreira V, Agapito A, et al. Non-VKA Oral Anticoagulants in Adult Congenital Heart Disease: a Single-Center Study. Cardiovasc Drugs Ther. 2023;37(6):1077–86.

18. Kartas A, Papazoglou AS, Moysidis DV, Despotopoulos S, Baroutidou A, Kosmidis D, et al. Use of apixaban in adults with congenital heart disease and atrial arrhythmias: The PROTECT-AR study. Int J Cardiol. 2024;406:131993.

19. Stalikas N, Doundoulakis I, Karagiannidis E, Bouras E, Kartas A, Frogoudaki A, et al. Non-Vitamin K Oral Anticoagulants in Adults with Congenital Heart Disease: A Systematic Review. J Clin Med. 2020;9(6).

20. Samarai D, Isma N, Lindstedt S, Hlebowicz J. Novel oral anticoagulant use in adults with congenital heart disease: a single-center experience report. Egypt Heart J. 2023;75(1):3.

21. Kazerouninia A, Georgekutty J, Kendsersky P, Byrne RD, Seto B, Chu PY, et al. A Multisite Retrospective Review of Direct Oral Anticoagulants Compared to Warfarin in Adult Fontan Patients. Cardiovasc Drugs Ther. 2023;37(3):519–27.

22. O’Shea MP, Yusuf A, Chao CJ, Ravi SN, karikalan s, Ashraf H, et al. RIGHT VENTRICULAR SYSTOLIC PRESSURE IS A SIGNIFICANT DETERMINANT OF EXCESS BLEEDING IN PEOPLE WITH NONVALVULAR ATRIAL FIBRILLATION TREATED WITH DABIGATRAN AND RIVAROXABAN COMPARED TO APIXABAN. JACC. 2024;83(13_Supplement):490-.

23. O’Shea MP, Yusuf A, Habib E, Ravi S, Karikalan SA, Chao CJ, et al. Echocardiographic Hemodynamics Correlate with Differences in DOAC-specific Bleeding and Stroke Rates in Non-Valvular Atrial Fibrillation. Research Square [PREPRINT]. 2024;1.

24. Dexter DD, Whisnant JP, Connolly DC, O’Fallon WM. The Association of Stroke and Coronary Heart Disease: A Population Study. Mayo Clinic Proceedings. 1987;62(12):1077–83.

25. Bhatia R, Sharma G, Patel C, Garg A, Roy A, Bali P, et al. Coronary Artery Disease in Patients with Ischemic Stroke and TIA. Journal of Stroke and Cerebrovascular Diseases. 2019;28(12).

26. Borre ED, Goode A, Raitz G, Shah B, Lowenstern A, Chatterjee R, et al. Predicting Thromboembolic and Bleeding Event Risk in Patients with Non-Valvular Atrial Fibrillation: A Systematic Review. Thromb Haemost. 2018;118(12):2171–87.

27. Chang G, Xie Q, Ma L, Hu K, Zhang Z, Mu G, et al. Accuracy of HAS-BLED and other bleeding risk assessment tools in predicting major bleeding events in atrial fibrillation: A network meta-analysis. J Thromb Haemost. 2020;18(4):791–801.

28. Hindricks G, Potpara T, Dagres N, Arbelo E, Bax JJ, Blomström-Lundqvist C, et al. 2020 ESC Guidelines for the diagnosis and management of atrial fibrillation developed in collaboration with the European Association for Cardio-Thoracic Surgery (EACTS): The Task Force for the diagnosis and management of atrial fibrillation of the European Society of Cardiology (ESC) Developed with the special contribution of the European Heart Rhythm Association (EHRA) of the ESC. European Heart Journal. 2020;42(5):373–498.

29. Joglar JA, Chung MK, Armbruster AL, Benjamin EJ, Chyou JY, Cronin EM, et al. 2023 ACC/AHA/ACCP/HRS Guideline for the Diagnosis and Management of Atrial Fibrillation: A Report of the American College of Cardiology/American Heart Association Joint Committee on Clinical Practice Guidelines. Circulation. 2024;149(1):e1–e156.

